# Molecular epidemiology of respiratory syncytial virus infections in Arizona, USA during the 2023-24 and 2024-25 seasons

**DOI:** 10.64898/2026.09.09.26362646

**Authors:** Steven C. Holland, LaRinda A. Holland, Tanner Porter, Veronica Boyle, Nivethitha Ramasamy, Michael White, Raquel Salgado, Martine Desulme, Lora Nordstrom, Mehal Patel, Jesus Estrada, Joanna Kramer, Efrem S. Lim, Neal Woodbury, Stacy White, Dave Engelthaler, Vel Murugan

**Author notes:** Authors contributed equally (Steven C. Holland, LaRinda A. Holland, Tanner Porter).

## Abstract

Respiratory syncytial virus infection is a common cause of hospitalizations in infants and elderly patients. With the increased availability of vaccines and therapeutic antibodies, surveilling the sequence landscape of viral genomes is important to increase awareness of potentially neutralizing mutations. During the 2023-24 and 2024-25 RSV seasons, we collected unpaired nasopharyngeal swab specimens from patients presenting to Arizona healthcare systems with symptoms of influenza-like illness. Using either tiled amplicon-based enrichment or a commercial oligo hybrid-enrichment panel and next generation sequencing, we obtained genome sequences for RSV and co-infecting viruses. We performed phylogenetic and genomic analyses to classify RSV genomes and detect mutations in the antigenic sites of the F protein and conserved central domain of the G protein. Phylogenetic analysis showed RSV-A strains belonged to the A.D. 1, A.D.2, A.D.3, and A.D. 5 subclades. RSV-B genomes belonged to the B.D.4.1, B.D.4.1.1, B.D.E.1, B.D.E.5, and B.D.E. 7 clades. We found 9 polymorphic sites located in the antigenic sites of the F protein in our RSV-A genomes and 18 polymorphic sites in our RSV-B genomes over the two-season surveillance period. We identified 7 polymorphic sites in our RSV-A genomes and 8 polymorphic sites in our RSV-B genomes that are in the conserved central domain of the G protein. We also discovered 3 (6.7%) and 27 (15.8%) incidences of viral coinfections in the 2023-24 and 2024-25 seasons, respectively. Viral genomic surveillance plays an important role in maintaining the efficacy of vaccine and therapeutic agents. Continued monitoring of circulating variants is an important component of public health.

## 1. Introduction

Respiratory syncytial virus (RSV) is a common cause of severe respiratory infection in young and elderly people. Infection may be particularly dangerous in children under 4 and adults older than 65 years of age, as it is a top contributor to hospitalization in these age groups [1]. With the recent approval of FDA-approved vaccines, genomic surveillance of circulating strains is important for vaccine and public health guidance.

RSV is a member of the *Pneumoviridae* family with a genome approximately 15,000 base pairs long, comprising 10 genes encoding 11 protein products. RSV is divided into 2 subtypes based upon G protein sequence: RSV-A and RSV-B. The G and F proteins play important roles in recognition and fusion of the virus to the host cell and may play additional roles in viral physiology [2].

The RSV F protein is of further interest due to its role as the primary immunologic target in host virus response as well as its biotechnological roles as vaccine antigen and therapeutic antibody target [3]. The commercially available ABRYSVO (Pfizer), AREXVY (GSK), and MRESVIA RSV (Moderna) vaccines use, as protein or mRNA precursor, a stabilized form of the F protein as their antigen [4]. Six antigenic sites of the F protein (Sites Ø-V) have been shown to be important binding sites for host and therapeutic antibodies [5]. Rare F protein mutations eliciting immune evasion have been discovered conferring resistance to clesrovimab, nirsevimab, and suptavumab therapeutic antibodies, among others [6-8].

The more variable G protein is also of evolutionary interest due to its high mutation rates and role in receptor recognition [9]. While highly variable, the RSV G protein sequences have a conserved central domain (CCD) housing a CX3C motif and heparin binding domain, through which the G protein binds the CX3C chemokine receptor and cell surface glycosaminoglycans, respectively [10].

Monitoring the current sequence landscape and evolutionary trends of these proteins, and the whole RSV genome, is important for maintaining and improving therapeutic efficacy. In this study we extend our previous 2022-23 RSV season analysis of RSV epidemiology in Arizona, USA, with the analysis of the 2023-24 and 2024-25 RSV seasons [11]. To assess the genomic RSV landscape, we recruited patients seeking care for influenza-like illnesses at healthcare facilities in Arizona. Viral whole genome sequencing was performed on RSV-positive nasopharyngeal swabs (NPS), followed by phylogenetic and genomic analyses.

## 2. Materials and Methods

### 2.1 Sample specimen recruitment

Patient specimens during the 2023-24 RSV season were obtained from two sources. The first sample cohort from the 2023-24 season (2023-24 Cohort #1) consisted of biobanked specimens obtained from patients seeking care for influenza-like illness at three Phoenix-area healthcare systems: Valley Wise Hospital, Phoenix Children’s Hospital, and Dignity Health Phoenix, which serve Maricopa County, Arizona, USA. At the time of the clinical visit, symptom questionnaires were completed by the patients or their guardians. During clinical evaluation, specimens were tested and confirmed negative for SARS-CoV-2 and influenza before biobanking. We tested 282 biobanked specimens for RSV using the TaqPath COVID-19, Flu A/B, RSV Combo Kit (Applied Biosystems, Waltham, MA). The qPCR assays and analysis were performed on a QuantStudio7 Flex Real-Time PCR Instrument (Applied Biosystems, Waltham, MA) and was performed per the manufacturer’s instructions. The 45 samples with C_t_ values less than 35 were selected for further viral whole genome sequencing and analysis. Respiratory specimens were stored in viral transport medium (VTM) at 4°C until nucleic acid extraction.

The second cohort for the 2023-24 season (2023-24 Cohort #2) consists of 75 biobanked samples obtained from healthcare systems servicing Coconino County, Arizona, USA. Samples were tested positive for RSV using standard-of-care respiratory pathogen testing performed at the healthcare system.

During the 2024-25 season, clinical specimens were obtained from patients seeking clinical care for influenza-like illness at Valleywise Health, which serves Maricopa County, Arizona, USA. Patient specimens tested positive for RSV using standard-of-care respiratory pathogen testing performed in the central clinical lab. Study participants provided informed written or verbal consent for study enrollment. Other samples were collected from clinical remnant samples, under a waiver of consent and HIPAA authorization.

This study was approved by Valleywise Health (Protocol ID 2024-056), Translational Genomics Institute (Protocol ID dengelthaler23-023ex) and Arizona State University (Protocol ID STUDY00011967) Institutional Review Board. Respiratory specimens were stored in (VTM) at 4°C until nucleic acid extraction.

### 2.2 Nucleic acid extraction, sequencing, analysis

For patient samples collected from 2023-24 Cohort #1 and all samples in 2024-25 season, nucleic acids were extracted using the MagMAX Viral/Pathogen II Nucleic Acid Isolation Kit (Applied Biosystems, Waltham, MA) and a Kingfisher Flex. The manufacturer’s MVP_2Wash_400_Flex program was run using up to 400 µL of NPS VTM was used as input and eluted into 200 µL of elution buffer.

NGS libraries for these patients were built using Illumina Respiratory Virus Oligo Panel v2 (Illumina, San Diego, CA), per the manufacturer’s protocol. Libraries were sequenced on an Illumina NextSeq2000 platform (Illumina, San Diego, CA) using 2×150-bp paired-end sequencing. After sequencing and demultiplexing, paired sequencing reads were trimmed of index and adapter sequences using trim galore version [12] (default parameters) and filtered from host reads using bbtools [13]. Reads were mapped to RSV-A and RSV-B reference genomes (GISAID accession nos. EPI_ISL_412866 and EPI_ISL_165399) using Burrows-Wheeler Aligner version 0.7.17-r1188 (default parameters) [14]. Consensus sequences were generated using SAMtools version 1.17 [15] (arguments -d 10, --min-MQ 20) at a minimum read depth ≥10. To detect co-infections, sequencing reads were also mapped to representative members of human adenovirus (22 sequences), human bocavirus (4 sequences), seasonal human coronaviruses (4 sequences), human metapneumavirus, human parechovirus (5 sequences), human parainfluenza viruses (5 sequences), human polyomaviruses (14 sequences), human influenza viruses (sequences comprising hemagglutinin and neuraminidases from influenza A, B, C), human rhinoviruses (3 sequences), and SARS-CoV-2, which were obtained from Genbank. Samples had to have ≥500 mapped reads and obtain ≥20% genome coverage at a minimum read depth ≥ 10 in order to be considered as a coinfection.

For samples collected in Coconino County (2023-24 Cohort #2), total nucleic acids were extracted using Zymo Quick-RNA Viral Kit R1035 (Zymo Research, Tustin, CA) and prepared for tiled amplicon sequencing [16]. Briefly, remnant UTM was added to Zymo DNA/RNA Shield with 1:1 ratio. 200 µL was then processed using the manufacturer’s specifications. Following extraction, 12 µL of nucleic acids underwent cDNA synthesis using 3 µL NEB LunaScript cDNA under the following protocol: 2 minutes at 25^°^C, 10 minutes at 55^°^C, and 95^°^C for 1 minute. After cDNA synthesis, RSV genomes were amplified using a previously published tiled amplicon primer set [17]. PCR amplicons were prepared for sequencing using a modified NEBNext Ultra II DNA Library kit. DNA concentrations were normalized for final pooling using NEBNext Library Quant Kit. Sequencing was performed on an Illumina NovaSeq 6000 or NextSeq 1000 using a p1 600-cycle kit. Sequencing reads were quality-filtered and analyzed as discussed above.

Phylogenetic analysis of RSV genomes was performed using Nextclade web software [18]. Statistical tests were performed using RStudio version 2025.09.1. Solvent exposed surface area measurements were made using the ShakeRupley algorithm (default parameters) within the Biopython library. The local antigenic site for a residue was defined as the contiguous upstream and downstream residues belonging to the antigenic site in which the mutated residue resides.

## 3. Results

### 3.1. Patient recruitment and demographics

To survey the genomic landscape of RSV in Arizona, USA, we collected surveillance samples from patients presenting with symptoms of influenza-like illness. Clinical diagnostic assays on nasopharyngeal swab samples confirmed RSV infection. During the 2023-24 RSV season, 120 total participants were recruited, with a median age of 11.25 years (IQR = 56.35 years) (**Figure 1A**). During the 2024 RSV season, 173 participants were included, with a median patient age of 7.0 (IQR = 41.58). Peak recruitment for the 2023-24 season occurred during MMWR weeks 1 and 6, corresponding to the week of December 31, 2023 and February 4, 2024, respectively (**FIGURE 1B**). Peak collection for the 2024-25 season occurred during MMWR week 6, corresponding to the week of February 2, 2025.

**Figure 1.**
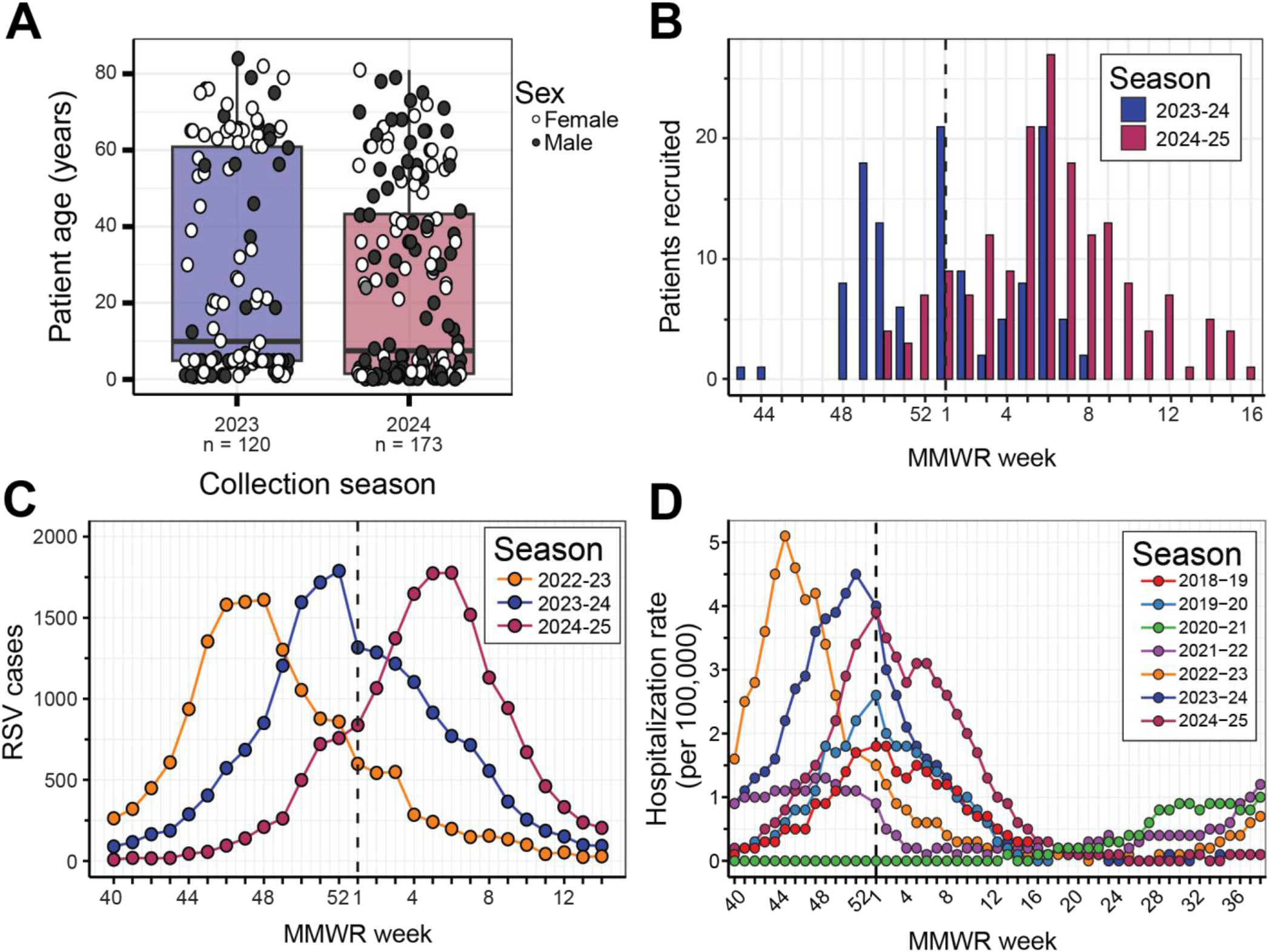
Cohort, Arizona, and USA RSV case frequencies. Patient age and sex demographics for clinical cohort. (**B**) Patient recruitment frequency for this study during the 2023-24 and 2024-25 RSV seasons. (**C**) RSV cases in Arizona, USA as measured by hospitalization cases. (**D**) RSV hospitalization rate in the USA.

Using publicly available hospitalization data published by the Arizona State Department of Public Health as a metric of clinical RSV activity [19], we found that our surveillance collection windows and peak collection periods were within state-wide peak RSV case activity (**FIGURE 1C**). We also observed that peak RSV activity in Arizona has occurred progressively later in the year but has not diminished in intensity over the 2022-23, 2023-24 and 2024-25 seasons. When we looked at national hospitalization rates reported by the CDC’s RSV Net, we observed that Arizona peak timings generally align with national trends but tend to occur towards the late shoulder of peak activity (**FIGURE 1D**). We observed that Arizona and national RSV peak timing has returned to historical norms after disruption of the 2020-21 season by the COVID-19 pandemic. National RSV activity appears to slightly decline over the 2022-2024 seasons but is still elevated compared to pre-COVID-19 levels.

### 3.2. Phylogenetic analysis of RSV genomes

Through whole genome sequencing of NPS samples, we identified 34 (29%) cases of RSV-A and 85 (71%) cases of RSV-B infection during the 2023-24 season. During the 2024-25 season, we identified 126 (81%) RSV-A and 29 (19%) RSV-B infections.

During the 2023-24 season, a 45-person cohort of patients were given symptom questionnaires during recruitment. Symptom presentations between persons with RSV-A and RSV-B infections were similar, with no significant statistical differences observed between the two viral subtypes for each symptom (**Table 1**; Fisher’s Exact test, *p* > 0.05).

**Table 1.**
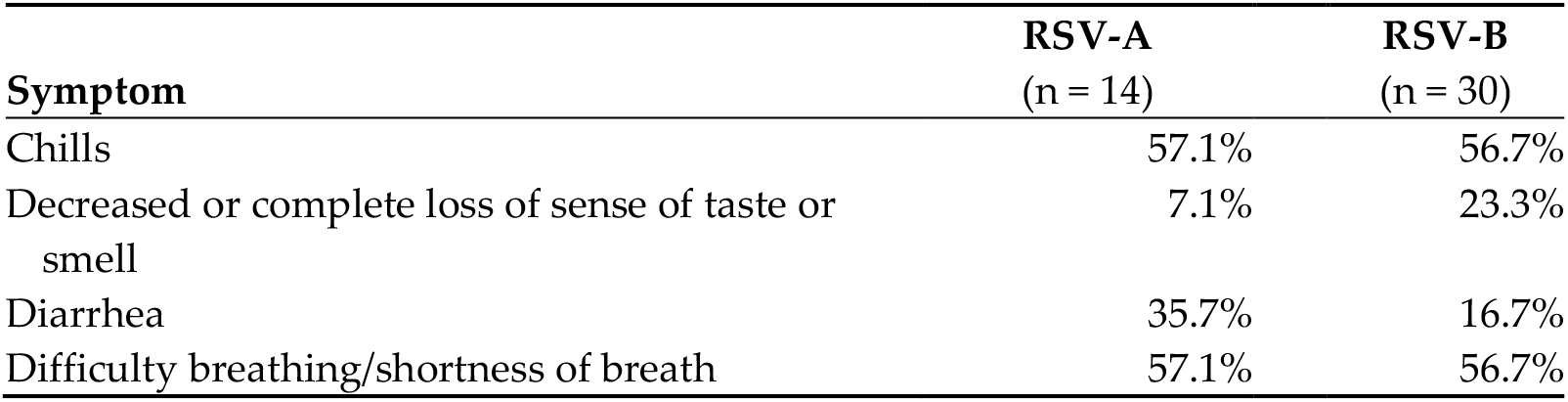
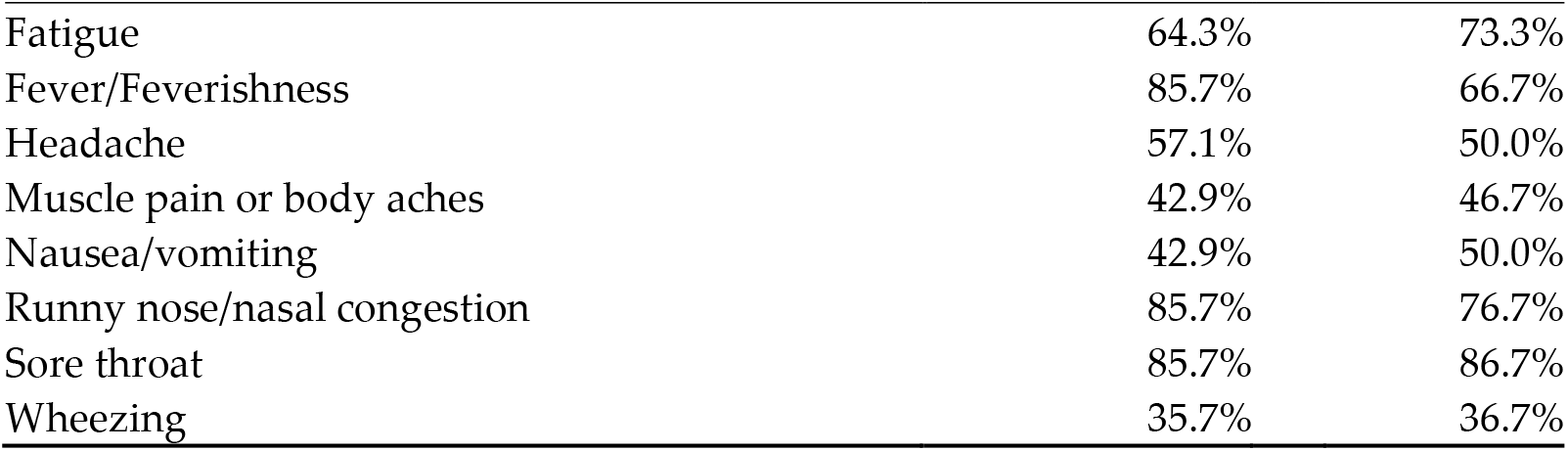
Percent of patients answering “Yes” to presentation of influenza-like symptoms.

| Symptom | RSV-A<br>(n = 14) | RSV-B<br>(n = 30) |
| --- | --- | --- |
| Chills | 57.1% | 56.7% |
| Decreased or complete loss of sense of taste or smell | 7.1% | 23.3% |
| Diarrhea | 35.7% | 16.7% |
| Difficulty breathing/shortness of breath | 57.1% | 56.7% |
| Fatigue | 64.3% | 73.3% |
| Fever/Feverishness | 85.7% | 66.7% |
| Headache | 57.1% | 50.0% |
| Muscle pain or body aches | 42.9% | 46.7% |
| Nausea/vomiting | 42.9% | 50.0% |
| Runny nose/nasal congestion | 85.7% | 76.7% |
| Sore throat | 85.7% | 86.7% |
| Wheezing | 35.7% | 36.7% |

Phylogenetic analysis of our moderate to high coverage (>60% genome coverage) RSV-A genomes revealed that specimens collected in Arizona during the 2023-24 and 2024-25 seasons belonged to the A.D.1, A.D.2, A.D.3, and A.D.5 clades (**Table 2**). In previous years, the A.D.5 comprised a majority of RSV-A genomes [11], this trend continued for the 2023-24 season, but A.D.5 became a minority of samples in the 2024-25 season.

**Table 2:** Phylogenetic classification of Arizona RSV genomes with moderate to high genome coverage (>60%) collected during the 2023-24 and 2024-25 seasons.

| RSV-A | 2023-24<br>(n = 33) | 2024-25<br>(n = 118) | RSV-B | 2023-24<br>(n = 82) | 2024-25<br>(n = 29) |
| --- | --- | --- | --- | --- | --- |
| A.D. | 0 (0%) | 5 (4%) | B.D.4.1 | 1 (2%) | 0 (0%) |
| A.D.1.4 | 0 (0%) | 3 (3%) | B.D.4.1.1 | 13 (16%) | 0 (0%) |
| A.D.1.5 | 3 (9%) | 11 (9%) | B.D.E.1 | 64 (78%) | 17 (59%) |
| A.D.1.6 | 2 (6%) | 11 (9%) | B.D.E.1.1 | 0 (0%) | 2 (7%) |
| A.D.1.8 | 2 (6%) | 0 (0%) | B.D.E.1.2 | 2 (2%) | 7 (24%) |
| A.D.1.9 | 1 (3%) | 12 (10%) | B.D.E.1.4 | 0 (0%) | 2 (7%) |
| A.D.1.11 | 0 (0%) | 2 (2%) | B.D.E.5 | 2 (2%) | 0 (0%) |
| A.D.2.1 | 1 (3%) | 0 (0%) | B.D.E.7 | 0 (0%) | 1 (3%) |
| A.D.3 | 5 (15%) | 41 (35%) |  |  |  |
| A.D.3.1 | 1 (3%) | 2 (2%) |  |  |  |
| A.D.3.2 | 3 (9%) | 2 (2%) |  |  |  |
| A.D.3.3 | 2 (6%) | 1 (1%) |  |  |  |
| A.D.3.9 | 0 (0%) | 6 (5%) |  |  |  |
| A.D.3.12 | 0 (0%) | 4 (3%) |  |  |  |
| A.D.5.1 | 4 (12%) | 2 (2%) |  |  |  |
| A.D.5.2 | 9 (27%) | 16 (14%) |  |  |  |

Phylogenetic analysis of our moderate to high coverage (>60% genome coverage) RSV-B genomes from samples collected in 2023-24 and 2024-25 showed that most genomes belonged to the B.D.E.1 clade and its sublineages (**Table 2**). We only observed sparse introductions of the B.D.4.1, B.D.E.1.1, B.D.E.5, and B.D.E.7 lineages during the two RSV seasons.

We next compared the phylogenetic placement of Nextclade reference sequences collected in the USA from 2022-2025 to our Arizona RSV-A and RSV-B genomes (**Figures 2C and 2D**). We observed that our RSV genomes generally clustered with USA reference sequences, indicating that viral genomes seeding Arizona infections were not unique to the region.

**Figure 2.**
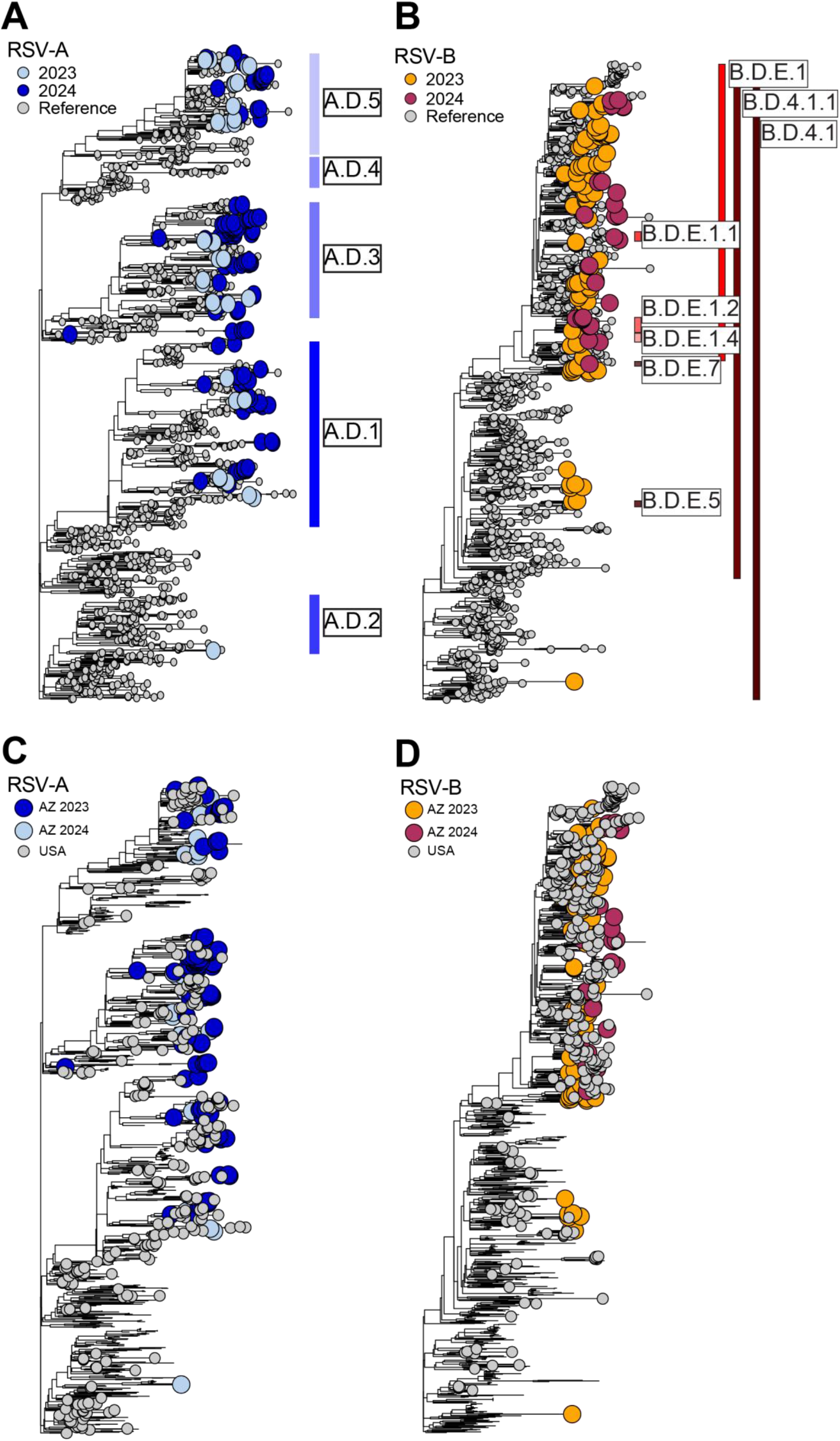
Phylogenetic trees of RSV-A (**A** and **C**) and RSV-B (**B** and **D**). Vertical bars show clade designations of clades containing genomes from this study. Top panels (**A** and **B**) show all Nextclade reference genomes as well as Arizona genomes colored by RSV season. Bottom panels (**C** and **D**) show Nextclade reference genomes collected from the USA as well as Arizona genomes.

### 3.3. Polymorphisms in antigenic sites of the F protein

Since the F protein is used as a target of many therapeutic antibodies and as a vaccine antigen, we explored polymorphisms in antigenic sites of the F protein from Arizona RSV samples (**Table 3**). For RSV-A samples, only three polymorphisms were found 2023-24 samples. Eight polymorphisms were detected in 2024-25 RSV-A samples. No RSV-A polymorphisms were found in high prevalence, with S276N being the most prevalent polymorphism detected in 12% of samples during 2023-24 and S377N, detected in 8% of 2024-25 samples.

**Table 3.**
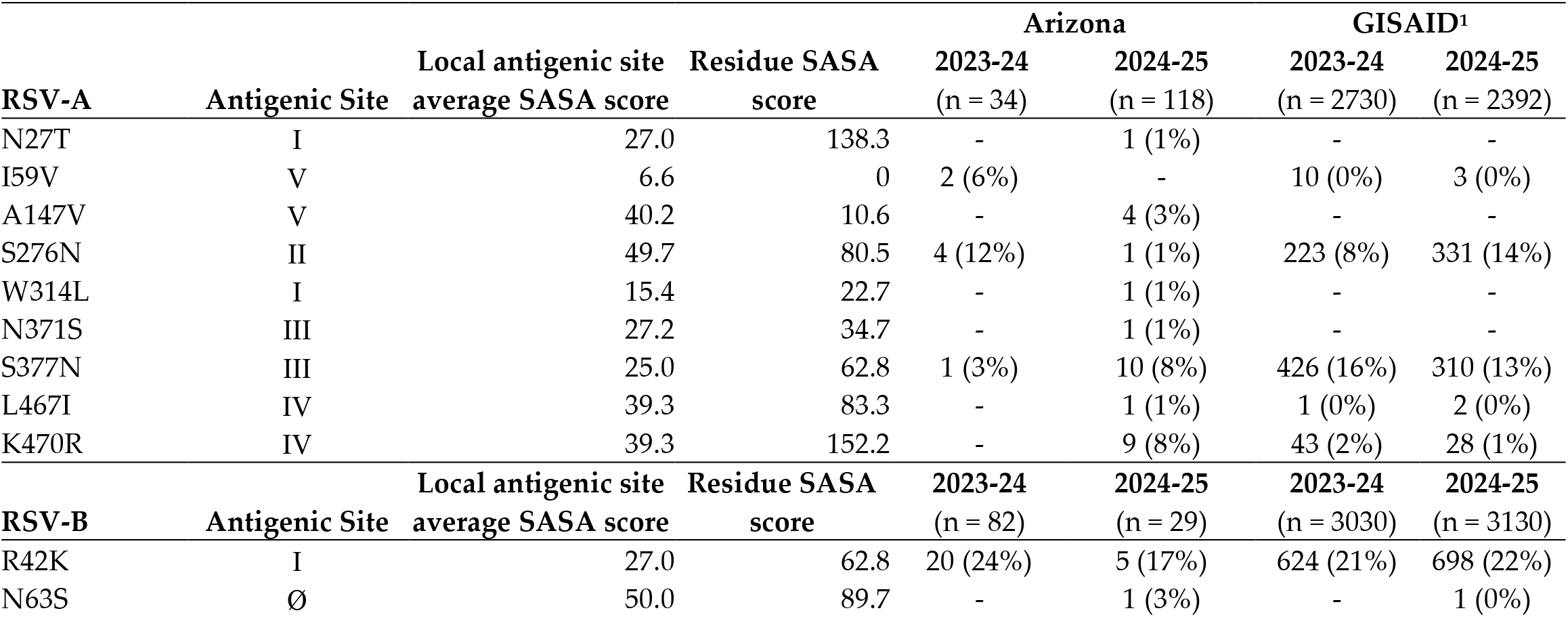
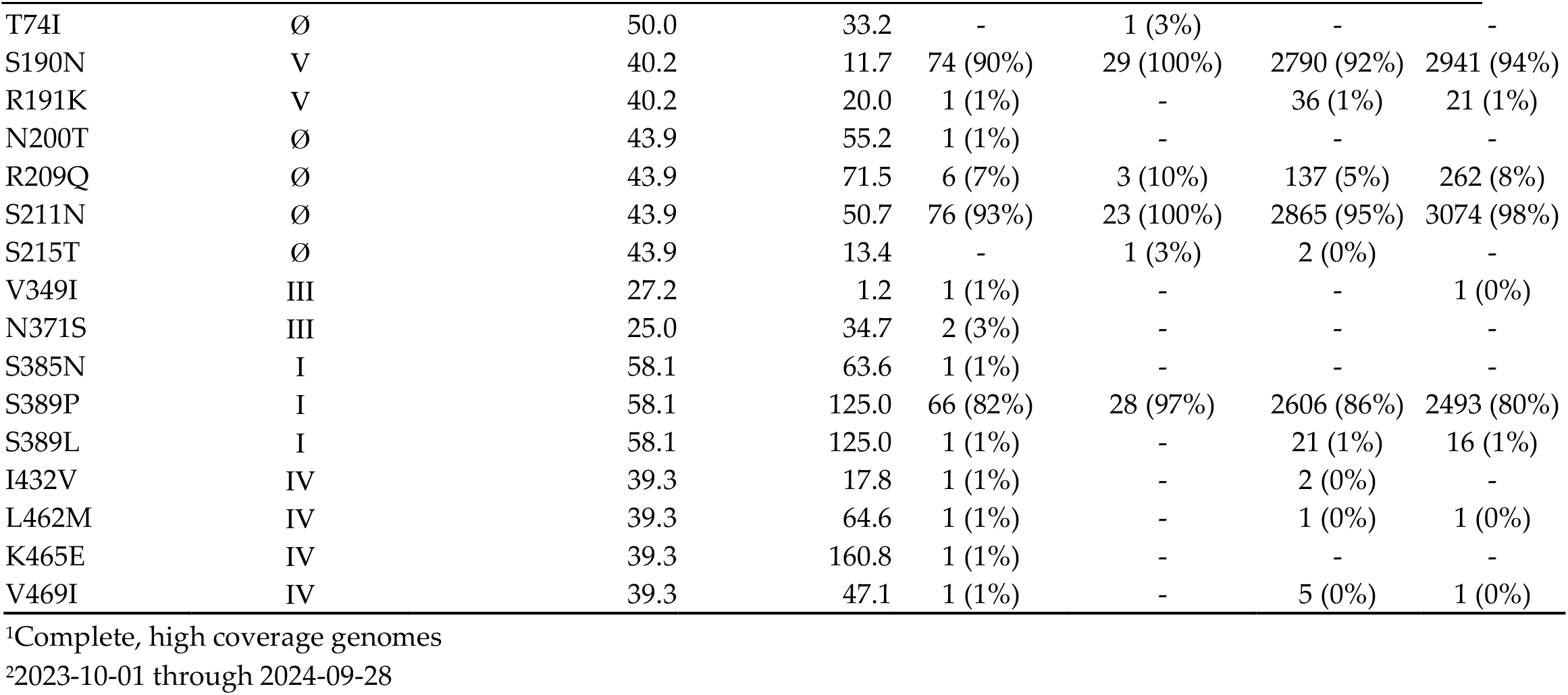
Discovered SNPs located in antigenic sites of the RSV F protein.

| RSV-A | Antigenic Site | Local antigenic site<br>average SASA score | Residue SASA<br>score | Arizona |  | GISAID<sup>1</sup> |  |
| --- | --- | --- | --- | --- | --- | --- | --- |
|  |  |  |  | 2023-24<br>(n = 34) | 2024-25<br>(n = 118) | 2023-24<br>(n = 2730) | 2024-25<br>(n = 2392) |
| N27T | I | 27.0 | 138.3 | - | 1 (1%) | - | - |
| I59V | V | 6.6 | 0 | 2 (6%) | - | 10 (0%) | 3 (0%) |
| A147V | V | 40.2 | 10.6 | - | 4 (3%) | - | - |
| S276N | II | 49.7 | 80.5 | 4 (12%) | 1 (1%) | 223 (8%) | 331 (14%) |
| W314L | I | 15.4 | 22.7 | - | 1 (1%) | - | - |
| N371S | III | 27.2 | 34.7 | - | 1 (1%) | - | - |
| S377N | III | 25.0 | 62.8 | 1 (3%) | 10 (8%) | 426 (16%) | 310 (13%) |
| L467I | IV | 39.3 | 83.3 | - | 1 (1%) | 1 (0%) | 2 (0%) |
| K470R | IV | 39.3 | 152.2 | - | 9 (8%) | 43 (2%) | 28 (1%) |
| RSV-B | Antigenic Site | Local antigenic site<br>average SASA score | Residue SASA<br>score |  |  |  |  |
|  |  |  |  | 2023-24<br>(n = 82) | 2024-25<br>(n = 29) | 2023-24<br>(n = 3030) | 2024-25<br>(n = 3130) |
| R42K | I | 27.0 | 62.8 | 20 (24%) | 5 (17%) | 624 (21%) | 698 (22%) |
| N63S | Ø | 50.0 | 89.7 | - | 1 (3%) | - | 1 (0%) |
| T74I | Ø | 50.0 | 33.2 | - | 1 (3%) | - | - |
| S190N | V | 40.2 | 11.7 | 74 (90%) | 29 (100%) | 2790 (92%) | 2941 (94%) |
| R191K | V | 40.2 | 20.0 | 1 (1%) | - | 36 (1%) | 21 (1%) |
| N200T | Ø | 43.9 | 55.2 | 1 (1%) | - | - | - |
| R209Q | Ø | 43.9 | 71.5 | 6 (7%) | 3 (10%) | 137 (5%) | 262 (8%) |
| S211N | Ø | 43.9 | 50.7 | 76 (93%) | 23 (100%) | 2865 (95%) | 3074 (98%) |
| S215T | Ø | 43.9 | 13.4 | - | 1 (3%) | 2 (0%) | - |
| V349I | III | 27.2 | 1.2 | 1 (1%) | - | - | 1 (0%) |
| N371S | III | 25.0 | 34.7 | 2 (3%) | - | - | - |
| S385N | I | 58.1 | 63.6 | 1 (1%) | - | - | - |
| S389P | I | 58.1 | 125.0 | 66 (82%) | 28 (97%) | 2606 (86%) | 2493 (80%) |
| S389L | I | 58.1 | 125.0 | 1 (1%) | - | 21 (1%) | 16 (1%) |
| I432V | IV | 39.3 | 17.8 | 1 (1%) | - | 2 (0%) | - |
| L462M | IV | 39.3 | 64.6 | 1 (1%) | - | 1 (0%) | 1 (0%) |
| K465E | IV | 39.3 | 160.8 | 1 (1%) | - | - | - |
| V469I | IV | 39.3 | 47.1 | 1 (1%) | - | 5 (0%) | 1 (0%) |

<sup>1</sup>Complete, high coverage genomes 391
<sup>2</sup>2023-10-01 through 2024-09-28 392

In the RSV-B genomes, we found 15 polymorphic sites in the 2023 season and 9 polymorphic sites in the 2024-25 season. In contrast to RSV-A polymorphisms, three RSV-B polymorphisms were found in high prevalence (>70% samples) across both 2023-24 and 2024-25.

We compared our Arizona genomes to high coverage RSV-A and RSV-B genomes in the GISAID database and observed that polymorphism abundances were concordant between sets. We did not observe any statistically significant differences in F protein polymorphism frequencies between GISAID and Arizona samples for any polymorphisms (Fisher Exact test, *p* > 0.05).

To explore if the discovered polymorphisms could affect antibody recognition, we mapped polymorphic sites to reference crystal structures (**FIGURE 3**). We observed that many polymorphisms occurred on surface exposed residues within the antigenic sites. To quantify the possible residue exposure, we calculated solvent accessible surface area (SASA) values for the reference residue as well as the average SASA value at the local antigenic site (**Table 3**). Most RSV-A (78%) and RSV-B (67%) polymorphic sites have SASA scores greater than the average SASA scores within their local antigenic site.

**Figure 3.**
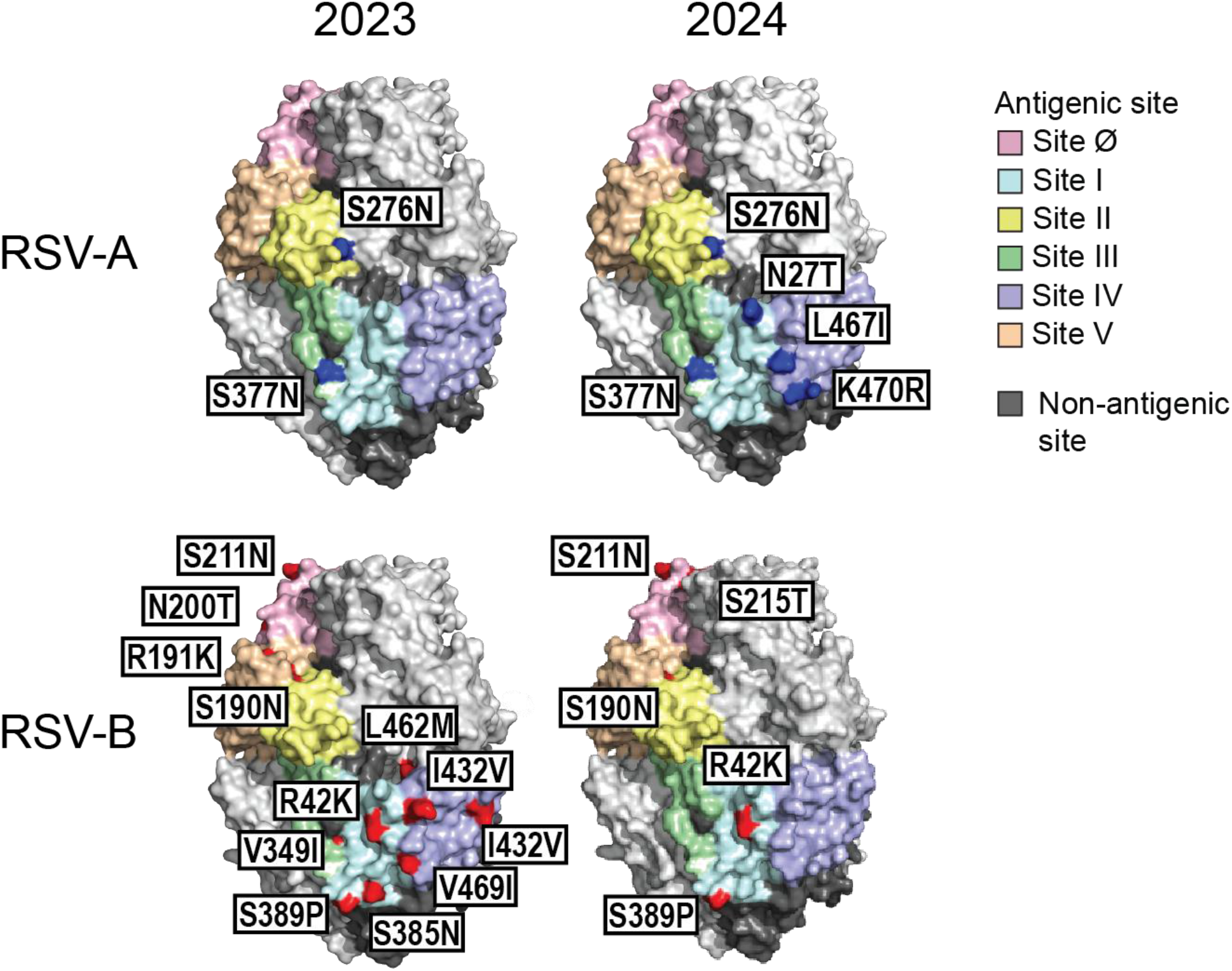
SNPs located within antigenic sites of the F protein. Top panels: RSV-A. Bottom panels: RSV-B. Note that some SNPs may be obscured by the protein orientation or may not be located on the protein surface. See **Table 2** for complete list.

### 3.4. Polymorphisms in the Conserved Central Domain of the G protein

Although the F protein is the major protein of immunological interest in RSV genomes, monitoring the evolutionary dynamics of the RSV G protein is important. When we looked at polymorphisms in the G protein CCD we observed seven polymorphisms in RSV-A samples and eight polymorphisms in RSV-B samples (**Table 4**). Of note, one polymorphism in the RSV-A samples, A184T, resides in the CX3C recognition motif of the G protein.

**Table 4.** Discovered SNPs located in the conserved central domain (∼aa 160-200) of the RSV G protein.

|  | <u>Arizona</u> |  | <u>GISAID</u> |  |
| --- | --- | --- | --- | --- |
|  | 2023-24<br>(n = 34) | 2024-25<br>(n = 118) | 2023-24<br>(n = 2730) | 2024-25<br>(n = 2392) |
| **RSV-A** |  |  |  |  |
| D162G | 1 | - | - | - |
| N178G | 11 (32%) | 31 (26%) | 768 (28%) | 1008 (42%) * |
| N178S | 1 (3%) | 20 (17%) | 4 (0%) | _* |
| A184T | - | 1 (1%) | 0 (0%) | 2 (0%) |
| N191S | - | 20 (17%) | 1 (0%) | 2 (0%) * |
| T200I | 1 (3%) | 1 (1%) | 9 (0%) | 6 (0%) |
| T200S | 1 (3%) | - | 37 (1%) | - |
| **RSV-B** |  |  |  |  |
|  | 2023-24<br>(n = 82) | 2024-25<br>(n = 29) | 2023-24<br>(n = 3030) | 2024-25<br>(n = 3130) |
| G175S | 1 (1%) | - | 9 (0%) | 4 (0%) |
| I187T | 1 (1%) | - | - | 2 (0%) |
| S189N | 1 (1%) | - | 11 (0%) | 30 (1%) |
| K193R | - | 1 (3%) | 5 (0%) | 2 (0%) |
| K195R | - | 2 (7%) | 3 (0%) | 16 (1%) |
| T197A | - | 3 (10%) | 6 (0%) | 51 (2%) |
| K199N | 4 (5%) | - | - | 4 (0%) |
| P200S | 1 (1%) | - | 8 (0%) | 1 (0%) |

\*Statistically significant frequency between Arizona and global GISAID samples. 394

Comparing our Arizona genomes to GISAID genomes revealed general concordance. We observed a statistically significant difference in the frequency of N178S and N191S mutations in our sequences (Fisher Exact test, p = 0.001). Manually inspecting G protein sequences revealed that the N178S and N191S mutations co-occurred in 19 of 20 instances during the 2024-25 season.

### 3.5. Respiratory virus coinfections with RSV

We also mapped sequencing reads to other respiratory viruses having probe targets on samples processed using a viral enrichment panel (2023: n = 45; 2024: n = 171). Using a minimum genome coverage threshold of 20%, we identified human adenovirus (HAdV), seasonal human coronavirus (HCoV), human metapneumovirus (HMPV), human parainfluenza (HPIV), human polyomavirus (HPyV), human influenza virus, human rhinovirus (HRV), and SARS-CoV-2 (SC2) coinfections within the patient samples (**FIGURE 4**). Three coinfections were identified in the 2023-24 cohort (6.7% incidence) and 27 coinfections were identified in the 2024-25 cohort (15.8% incidence). Three multiple virus (3+) co-infections were detected and were comprised of RSV/HAdV/HCoV, RSV/HCoV/HPyV, and RSV/HCoV/HMPV infections. Sequence read mapping to reference strains revealed multiple subtypes of HAdV, HCoV, HPyV, and influenza viruses. There was no statistically significant difference in patient ages between patients with and without coinfections for the 2023-24 and 2024-25 seasons (Student’s t-Test, *p* > 0.05).

**Figure 4.**
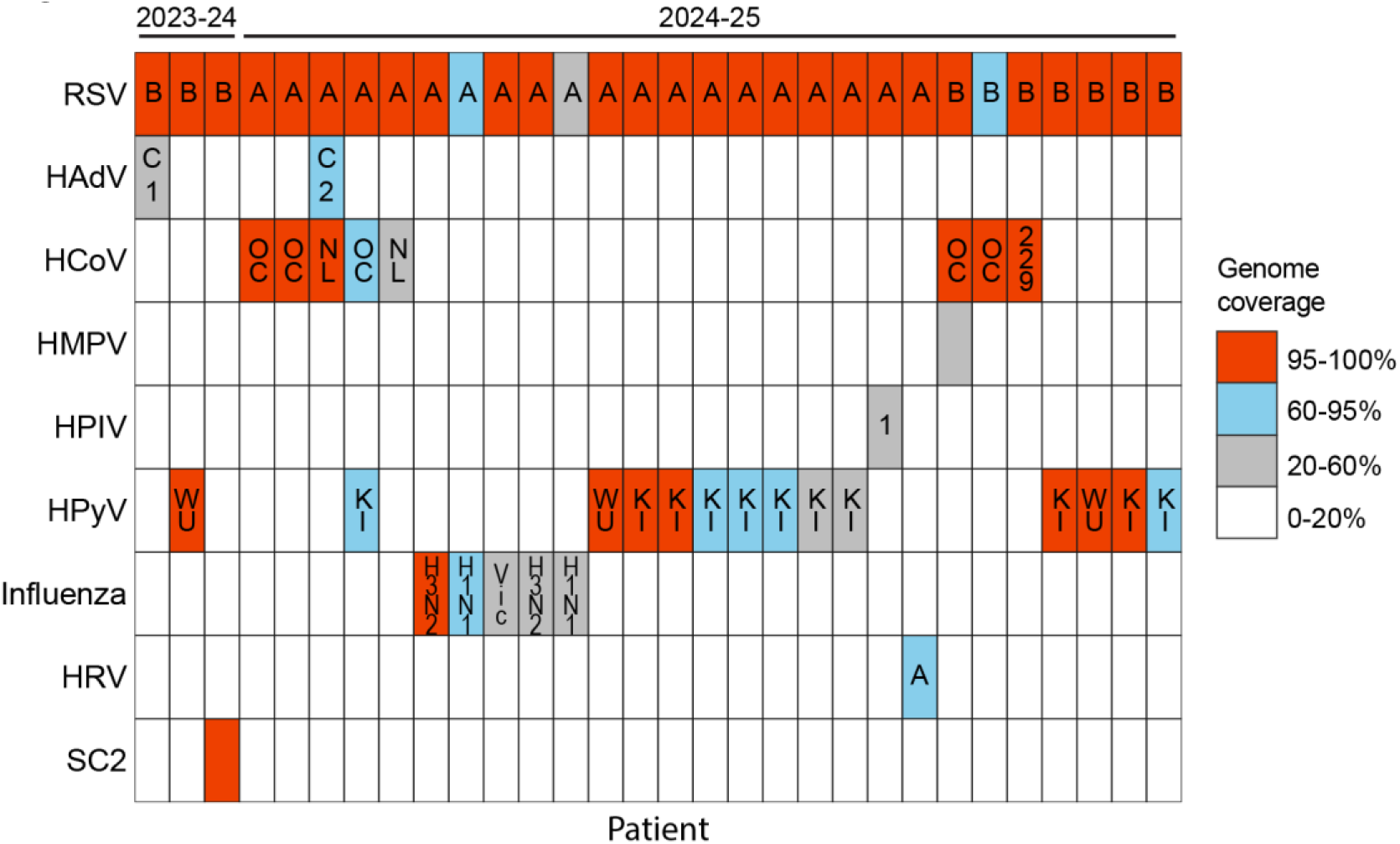
Viral coinfections found in RSV samples. Coloration depicts percent genome recovered. Letters and numbers indicate viral subtypes when applicable. Subtypes and abbreviations: HAdV: human adenoviruses C1 & C2, HCoV: human coronaviruses OC43 & NL63 & 229E, HMPV: human metapneumovirus, HPIV: human parainfluenza virus 1, HPyV: human polyomaviruses WU polyomavirus & KI polyomavirus, Influenza A/H3N2 & H1N1 & B/Victoria, HRV: human rhinovirus A, SC2: SARS-CoV-2.

## 4. Discussion

In this study, we sequenced nasopharyngeal swabs from 293 patients and obtained 262 RSV genomes with coverage >60% during the 2023-24 and 2024-25 RSV seasons in Arizona, USA. We identified RSV-A and RSV-B genomes in our cohorts, with the majority subtype frequency swapping between seasons. We identified 9 polymorphisms in antigenic sites of the F protein in RSV-A as well as 18 polymorphisms within the antigenic sites of RSV-B genomes. We also observed 7 and 8 polymorphisms in the CCD of the G protein in RSV-A and RSV-B genomes, respectively. We also identified 30 potential respiratory virus co-infections with RSV over the two-year surveillance.

Peak RSV activity in Arizona has occurred progressively later in the respiratory virus season after its disruption due to the COVID-19/SARS-CoV-2 pandemic [20]. Nationally, the 2023-24 and 2024-25 RSV seasons were also slightly lower in activity than the 2022-23 season but remained steady in Arizona. Other studies reporting national trends suggest lower activity is in part due to the protective effects of RSV vaccine and nirsevimab administration, as hospitalization decreases were largest in cohorts receiving interventions [21]. Arizona’s immunization coverage of 24.6% in infants [22] and 11.8% in 60+ year old adults [23] is near median national values of 31.1% and 11.4%, respectively. However, our multi-year genomic surveillance supports behavioral or interventional causes to explain changes in RSV activity, since no notable novel RSV variants were observed, nor did we observe dominance of a particular variant over our multi-year surveillance.

We observed a notable difference in RSV-A and RSV-B abundances between the 2023-24 and 2024-25 seasons. In 2023-24, RSV-A comprised only 29% of our recovered genomes, whereas it comprised 81% of recovered genomes in 2024-25. In the 2022-23 season, we previously observed that RSV-A was the dominant circulating subtype (79%) in Arizona [11]. RSV subtype abundance does change from year to year, although RSV-A is slightly more often dominant [24]. Subtype abundance appears to change stochastically, as there is rarely evidence of more virulent strains evolving. Our phylogenetic analyses are consistent with this pattern, as introduced variants were located within already established clades and evolutionary divergences are consistent with historical trends.

We identified 27 total polymorphisms within antigenic sites of the F protein. A literature search for our annotated polymorphisms revealed no evidence of these sites significantly affecting virus physiology or pathogenicity. Similarly, we observed 15 total polymorphisms in the CCD region of the G protein. Notably, the A184T polymorphism resides in the CX3C binding site. We were unable to find an annotated effect of this and other G protein mutations.

Multiple respiratory viruses co-circulate with RSV during the respiratory virus season. In our study, we found 30 patients with possible respiratory virus co-infections. Most RSV co-infections were with human polyomaviruses WUPyV and KIPyV (14/30 coinfections). These polyomaviruses have often been observed to co-infect patients with respiratory infections, and no known pathogenic effects have been described [25]. We also found seasonal coronaviruses OC43, NL63, and 229E in patient samples. We found OC43 in highest prevalence, which has been reported in other studies [26, 27]. The observed HAdV-C2, HPIV-1, Influenza H3N2/H1N1/Vic, and HRV-A viruses are all common respiratory infection subtypes found to co-circulate with RSV [28-31]. Since qPCR and many other clinical assays are limited in their multiplexing, the current rate of RSV co-infection may be underestimated. As multi-target enrichment or non-targeted metagenomic analyses become more prevalent, our understanding of the incidence of RSV co-infection will become more refined.

There are limitations to this study. Because a large portion of sample recruitment was performed at a children’s hospital, our surveillance cohort skews towards young participant ages, as well as patients that felt infection symptoms warranted seeking medical attention. Median ages in our surveillance cohorts were 11.25 and 7.0 years for the 2023-24 and 2024-25 seasons, respectively. As pediatric RSV respiratory infections vary in subtype preference and presentation compared to adult infections [1], caution should be utilized when generalizing our results or comparing them to other studies.

RSV has become increasingly recognized as an important contributor to acute respiratory illnesses. As RSV continues to evolve, surveillance of the RSV F protein for polymorphisms affecting vaccine and therapeutic efficacy is of high importance. With the ability to survey other RSV genes and detect novel variants of increase virulence, NGS can play an important role in epidemiological monitoring. NGS can also detect mutations that may confer resistance to therapeutics and survey for other respiratory pathogens present in clinical samples. These factors may play a role in the uptake of NGS for diagnostic applications, as sequencing prices continue to lower and diagnostic NGS becomes increasingly available.

## Data Availability

All data produced in the present study are available upon reasonable request to the authors

## Author Contributions

Conceptualization, S.C.H., L.N., E.S.L., D.E., and V.M.; methodology, S.C.H., T.P; software, S.C.H.; validation, S.C.H., T.P., and L.A.H.; formal analysis, S.C.H., T.P; investigation, S.C.H., L.A.H., T.P., V.B., N.R., M.W., R.S., S.W., and M.D.; resources, L.A.H., J.K., M.P., J.E., and L.N.; data curation, S.C.H.; writing—original draft preparation, S.C.H.; writing - review and editing, D.E., and V.M.; visualization, S.C.H.; supervision, M.W., J.K., L.N., and V.M.; project administration, L.N., E.S.L., N.W., D.E., and V.M.; funding acquisition, N.W., D.E., and V.M. All authors have read and agreed to the published version of the manuscript.

## Institutional Review Board Statement

The study was conducted in accordance with the Declaration of Helsinki, and approved by the Institutional Review Boards of Valleywise Health (Protocol ID 2024-056, approved on 13 August 2024), Translational Genomics Institute (Protocol ID dengelthaler23-023ex, approved on 20 December 2023) and Arizona State University (Protocol ID STUDY00011967, approved 20 May 2020).

## Informed Consent Statement

Informed consent or a waiver of consent and HIPAA authorization was obtained for collection of all specimens analyzed in the study.

## Data Availability Statement

RSV consensus genomes have been submitted to the GISAID EpiRSV database and can be accessed using EPISET EPI_SET_260813mr. Consensus genomes of viral coinfections have been submitted to the GISAID EpiFlu (Influenza: EPI_ISL_20499901 & EPI_ISL_20499899) and Genbank databases (HCoV, HMPV, HRV: PZ38588-PZ738597; HAdV, HPyV: PZ744101-PZ744113; SC2: PZ736741).

## Acknowledgments

We would like to thank the ASU Sequencing Core for their assistance in next generation sequencing. We also thank Allison Glazer and Parker Montfort from Translational Genomics Institute for their help in sample handling. We also recognize and thank the sequence authors and contributors of the GISAID and Genbank databases.

## Abbreviations

The following abbreviations are used in this manuscript:

HadV: Human adenovirus
HcoV: Human coronavirus
HMPV: Human metapneumovirus
HPIV: Human parainfluenza virus
HpyV: Human polyomavirus
HRV: Human rhinovirus
NGS: Next generation sequencing RSV Respiratory syncytial virus
SC2: SARS-CoV-2; severe acute respiratory syndrome coronavirus 2

